# The Use of Adjuvant Therapy in Preventing Progression to Severe Pneumonia in Patients with Coronavirus Disease 2019: A Multicenter Data Analysis

**DOI:** 10.1101/2020.04.08.20057539

**Authors:** Zhichao Feng, Jennifer Li, Shanhu Yao, Qizhi Yu, Wenming Zhou, Xiaowen Mao, Huiling Li, Wendi Kang, Xin Ouyang, Ji Mei, Qiuhua Zeng, Jincai Liu, Xiaoqian Ma, Pengfei Rong, Wei Wang

## Abstract

**IMPORTANCE:** Coronavirus disease 2019 (COVID-19) is a global pandemic associated with high mortality and effective treatment to prevent clinical deterioration to severe pneumonia has not yet been well clarified.

**OBJECTIVE:** To investigate the role of several adjuvant treatments in preventing severe pneumonia in patients with COVID-19.

**DESIGN, SETTING, AND PARTICIPANTS:** Multicenter, retrospective cohort study of 564 consecutively hospitalized patients with confirmed COVID-19 at Third Xiangya Hospital of Central South University, Changsha Public Health Treatment Center, First Hospital of Yueyang, Junshan People’s Hospital of Yueyang, Central Hospital of Shaoyang, Central Hospital of Xiangtan, Second Hospital of Changde, Central Hospital of Loudi, and First Affiliated Hospital of University of South China in Hunan province from January 17, 2020 to February 28, 2020; The final date of follow-up was March 15, 2020.

**EXPOSURES:** Nonspecific antivirals (arbidol, lopinavir/ritonavir, and interferon α), antihypertensives, and chloroquine.

**MAIN OUTCOMES AND MEASURES:** The development of severe COVID-19 pneumonia; Demographic, epidemiological, clinical, laboratory, radiological, and treatment data were collected and analyzed.

**RESULTS:** Of 564 patients, the median age was 47 years (interquartile range, 36-58 years), and 284 (50.4%) patients were men. Sixty-nine patients (12.2%) developed severe pneumonia. Patients who developed severe pneumonia were older (median age of 59 and 45 years, respectively), and more patients had comorbidities including hypertension (30.4% and 12.3%, respectively), diabetes (17.4% and 6.7%, respectively), and cardiovascular disease (8.7% and 3.2%, respectively) and presented with fever (84.1% and 60.4%, respectively) and shortness of breath (10.1% and 3.8%, respectively) compared with those who did not. Nonspecific antiviral therapy did not prevent clinical progression to severe pneumonia, although fewer hypertensive patients on angiotensin-converting enzyme inhibitors or angiotensin-receptor blockers (ACEI/ARB) therapy developed severe pneumonia in contrast with those on non-ACEI/ARB antihypertensive therapy (1 of 16 [6.3%] patients and 16 of 49 [32.7%] patients, respectively [difference, 26.4%; 95% CI, 1.5% to 41.3%]). Multivariate logistic regression analysis showed that hypertension without receiving ACEI/ARB therapy was an independent risk factor (odds ratio [OR], 2.07; 95% CI, 1.07 to 4.00) for developing severe pneumonia irrespective of age. Besides, none of patients treated with chloroquine developed severe pneumonia, though without significance (difference, 12.0%; 95% CI, −3.5% to 30.0%) by propensity score matching.

**CONCLUSIONS AND RELEVANCE:** Hypertensive patients on ACEI or ARB may be protective from severe pneumonia in COVID-19 and hence these therapies should not be ceased unless there is a strong indication or further epidemiological evidence. Though none of the current antiviral and immunoregulation therapy showed benefit in preventing COVID-19 progression, chloroquine deserved further investigation.

**KEYPOINTS:** *Question:* Does the use of adjuvant therapy reduce progression to severe pneumonia in patients with coronavirus disease 2019 (COVID-19)?

*Findings:* In this retrospective, observational cohort study involving 564 patients with confirmed COVID-19, hypertension was an independent risk factor for progression to severe pneumonia irrespective of age and those on angiotensin-converting enzyme inhibitor (ACEI) or angiotensin receptor blocker (ARB) therapy were less likely to develop severe COVID-19 pneumonia, while nonspecific antivirals or chloroquine did not have significant impact on clinical progression.

*Meaning:* Hypertensive patients with COVID-19 should not have ACEI or ARB ceased, unless there is a strong indication or further epidemiological evidence, given its potential protective effects.

## INTRODUCTION

The world has witnessed the rapid escalation of 2019 coronavirus disease (COVID-19), caused by severe acute respiratory syndrome coronavirus 2 (SARS-CoV-2), now becoming a global pandemic.^1^ While many patients may be asymptomatic or experience only mild symptoms, reports so far have identified the elderly and patients with chronic comorbidities such as hypertension, diabetes, and cardiovascular disease to be at higher risk of progression to severe COVID-19 pneumonia, characterized by acute respiratory distress syndrome (ARDS) and/or multiorgan failure. ^2-4^ These severe patients require more intensive medical resource utilization and have worse prognosis, with a case fatality rate about 20 times higher than that of non-severe patients. ^5,6^ While the implementation of effective prevention measures is essential, there clearly exists an urgent need for interventions to modulate disease progression in the face of an increasing number of infected individuals, which have overwhelmed the capacity of hospital systems in some countries.

While there is no specific therapy for COVID-19, current progress with investigational therapies includes the development and trial of a coronavirus vaccine and interrogation of several orphan drugs in randomized control trials (RCTs).^7^ This includes remdesivir, which has garnered the most interest in an ongoing adaptive RCT and other nucleotide analogues including ritonavir, favipiravir and galidesvir.^8^ A recently published open-label RCT showed no benefit of the protease inhibitors lopinavir/ritonavir in reducing clinical severity or mortality associated with COVID-19, despite in vitro evidence of activity against SARS-CoV-2.^9^ Other drugs of interest include antiviral therapy (pegylated interferon-alpha, arbidol) and immunomodulators (hydroxychloroquine) are also under investigation and results are eagerly monitored.^10,11^

Inhibition of the spike glycoprotein (S-protein) may also be a viable target and has recently garnered increasing interest - particularly the potential interactions with angiotensinogen converting enzyme 2 (ACE2). It is proposed that the binding of the SARS-CoV-2 S-protein to the functional receptor ACE2 results in proteolytic cleavage by type II transmembrane serine proteases (TMPRSS2).^12-14^ There has been considerable interest and debate regarding the risk of severe COVID-19 pneumonia in patients on angiotensinogen converting enzyme inhibitors (ACEI) or angiotensin receptor blocker (ARB) therapy, presumably through the modulation of ACE2 expression.^4,15,16^ The role of ACEI and ARB in COVID-19 infection remains controversial – as the data is not adjusted for confounding factors of age and other comorbidities to definitively show that ACEI or ARB are independently associated with worse outcomes – and of course, this does not show causality either.^17^ Hence, various independent medical societies have put out position statements that recognize the potential risks but given the current quality of evidence, recommend against inappropriate cessation or switch from ACEI/ARB therapy for hypertension, given its proven benefits.^18,19^

Therefore, the objective of this study was to investigate the role of several adjuvant treatments in preventing severe pneumonia in COVID-19 patients from seven cities in Hunan province, China.

## METHODS

### Study Design and Participants

The Institutional Review Board of Third Xiangya Hospital of Central South University approved our study and waived informed consent for the retrospective nature of this study. All consecutive adult patients with confirmed COVID-19 who were treated at Third Xiangya Hospital of Central South University, Changsha Public Health Treatment Center, First Hospital of Yueyang, Junshan People’s Hospital of Yueyang, Central Hospital of Shaoyang, Central Hospital of Xiangtan, Second Hospital of Changde, Central Hospital of Loudi, and First Affiliated Hospital of University of South China from January 17, 2020 to February 28, 2020 were enrolled. The diagnosis of COVID-19 was established based on the World Health Organization (WHO) interim guidance, and a confirmed case was defined as a positive result to high-throughput sequencing or real-time reverse transcription-polymerase chain reaction (RT-PCR) assay for nasal and pharyngeal swab specimens. Patients without available clinical or imaging data were excluded. The clinical outcomes including severe pneumonia development, discharge, and death were monitored up to March 15, 2020.

### Clinical and imaging data collection

We extracted the demographic, exposure history, comorbidities, clinical symptoms, and laboratory data on admission from electronic medical records. Medication history among individuals with hypertension was recorded, including ACEI/ARB, calcium-antagonists, β-blocker, and diuretic agents. All laboratory tests were performed according to the clinical care needs of the patient during hospitalization, including a complete blood count, assessment of liver and renal function, coagulation testing, and measures of electrolytes, creatine kinase, lactate dehydrogenase, procalcitonin, and C-reactive protein. The use of antiviral therapy (arbidol, lopinavir/ritonavir, and interferon α) and chloroquine were obtained from the medication administration record. The severity degree of COVID-19 at the time of admission is defined as follows: (1) mild type, with fever or respiratory tract symptoms but without radiological pneumonia. (2) moderate type, with fever, respiratory tract symptoms, and radiological evidence of pneumonia. (3) severe type, with one of the following: a) respiratory distress (respiratory rate ≥ 30 beats/min); b) hypoxia (oxygen saturation ≤ 93% in the resting state); c) hypoxemia (arterial blood oxygen partial pressure/oxygen concentration ≤ 300mmHg). (4) critical type, with one of the following: a) respiratory failure requiring mechanical ventilation; b) shock; c) intensive care unit (ICU) admission is required for combined other organs failure. In this study, severe COVID-19 pneumonia broadly included severe and critical types.

The CT images were acquired on admission and at intervals of 3-5 days and were reviewed independently by two experienced radiologists. Typical CT findings for COVID-19 were defined as peripherally distributed multifocal ground-glass opacities (GGOs) with patchy consolidations and posterior part or lower lobe involvement predilection.^20^ Each of the five lung lobes was reviewed for GGO and consolidation. The lesions extent within each lung lobe was semi-quantitatively evaluated by scoring from 0 to 5 based on the degree of involvement: score 0, none involvement; score 1, ≤ 5% involvement; score 2, 6%∼25% involvement; score 3, 26%∼50% involvement; score 4, 51%∼75% involvement; score 5, > 75% involvement. The total score was calculated by summing up scores of all five lobes to provide a lung CT score ranging from 0 to 25.^21^ The mean value of lung CT scores evaluated by the two radiologists was used for analysis.

### Statistical Analysis

Our primary outcome of analysis was progression to severe COVID-19 pneumonia. Continuous variables are presented as the median and interquartile range (IQR), and categorical variables are presented as frequency and percentage. Differences between groups were analyzed using Student’s t-test or Mann-Whitney U test for quantitative variables according to the normal distribution and Chi-square test or Fisher’s exact test for categorical variables. Meanwhile, the differences in distributions were also reported using differences with 95% confidence intervals (CIs). Univariate and multivariate logistic regression with backward stepwise selection based on likelihood ratio was used to identify the risk factors for the development of severe pneumonia, and the corresponding odds ratios (ORs) and 95% CIs were determined. We also employed the propensity score matching (PSM) with a 1:1 ratio to minimize selection bias and adjust for the imbalance between patients with or without chloroquine therapy. A one-to-one nearest-neighbour matching algorithm with an optimal calliper of 0.2 without replacement was used to generate pairs of patients. The analyses regarding different factors were based on non-missing data, and missing data were not imputed. All statistical analyses were performed using SPSS statistics software (version 22.0, IBM SPSS Inc, Chicago, IL, USA). A two-sided *P* value of less than 0.05 considered to be statistically significant.

## RESULTS

### Patient Characteristics

The study population included 564 adult patients with confirmed COVID-19 from nine hospitals in seven cities (Figure 1), and their clinical characteristics on admission are presented in Table 1. The median age of patients was 47 years (IQR, 36-58; range, 19-84 years), and 284 (50.4%) were men. 132 (23.4%) patients had comorbidities, including hypertension (n = 82 [14.5%]), diabetes (n = 45 [8.0%]), cardiovascular disease (n = 22 [3.9%]), and chronic obstructive pulmonary disease (COPD, n = 16 [2.8%]). The most commonly self-reported symptoms at onset of illness were fever (n = 357 [63.3%]), cough (n = 323 [57.2%]), fatigue (n = 87 [15.4%]), sputum production (n = 51 [9.0%]), anorexia (n = 26 [4.6%]), and shortness of breath (n = 26 [4.6%]). A total of 516 (91.5%) patients had typical abnormal findings on chest CT (Supplementary Figure 1), with a median lung CT score of 6 (IQR, 4-11; range, 0-25).

**Table 1.** Clinical characteristics of 564 COVID-19 patients according to disease severity

| Characteristics | Total (n =564) | Severe (n = 69) | Non-Severe (n = 495) | Difference (95% CI) | *P* |
| --- | --- | --- | --- | --- | --- |
| Age (years) | 47 (36 to 58) | 59 (51 to 66) | 45 (35 to 56) | -13 (-16 to -9) | **< 0.001** |
| < 50 | 317 (56.2%) | 16 (23.2%) | 301 (60.8%) |  |  |
| 50~65 | 160 (28.4%) | 31 (44.9%) | 129 (26.1%) |  |  |
| ≥ 65 | 87 (15.4%) | 22 (31.9%) | 65 (13.1%) |  |  |
| Male gender | 284 (50.4%) | 39 (56.5%) | 245 (49.5%) | -7.0 (-18.9 to 5.5) | 0.274 |
| Exposure within 14 days |  |  |  | -7.1 (-19.3 to 5.2) | 0.267 |
| recently been to Wuhan | 251 (44.5%) | 35 (50.7%) | 216 (43.6%) |  |  |
| contacted with people from Wuhan or local infected patients | 313 (55.5%) | 34 (49.3%) | 279 (56.4%) |  |  |
| Smoking history | 47 (8.3%) | 9 (13.0%) | 38 (7.7%) | -5.4 (-15.5 to 1.2) | 0.131 |
| Comorbidity |  |  |  |  |  |
| Any | 132 (23.4%) | 37 (53.6%) | 95 (19.2%) | -34.4 (-46.2 to -22.2) | **< 0.001** |
| Hypertension | 82 (14.5%) | 21 (30.4%) | 61 (12.3%) | -18.1 (-30.1 to -8.0) | **< 0.001** |
| Diabetes | 45 (8.0%) | 12 (17.4%) | 33 (6.7%) | -10.7 (-21.5 to -3.1) | **0.002** |
| Cardiovascular disease | 22 (3.9%) | 6 (8.7%) | 16 (3.2%) | -5.5 (-14.6 to -0.4) | **0.028** |
| COPD | 16 (2.8%) | 9 (13.0%) | 7 (1.4%) | -11.6 (-21.6 to -5.4) | **< 0.001** |
| Cerebrovascular disease | 5 (0.9%) | 0 (0) | 5 (1.0%) | 1.0 (-4.3 to 2.3) | 0.402 |
| Chronic renal disease | 3 (0.5%) | 2 (2.9%) | 1 (0.2%) | -2.7 (-9.8 to 0.4) | **0.004** |
| Hepatitis B/C infection | 9 (1.6%) | 3 (4.4%) | 6 (1.2%) | -3.1 (-10.8 to 0.1) | 0.051 |
| Malignancy | 4 (0.7%) | 1 (1.5%) | 3 (0.6%) | -0.8 (-7.2 to 0.8) | 0.434 |
| Immunodeficiency | 1 (0.2%) | 0 (0) | 1 (0.2%) | 0.2 (-5.1 to 1.1) | 0.709 |
| Symptoms or signs |  |  |  |  |  |
| Fever | 357 (63.3%) | 58 (84.1%) | 299 (60.4%) | -23.7 (-31.7 to -12.4) | **< 0.001** |
| Cough | 323 (57.2%) | 37 (53.6%) | 286 (57.8%) | 4.2 (-7.9 to 16.6) | 0.513 |
| Fatigue | 87 (15.4%) | 8 (11.6%) | 79 (16.0%) | 4.4 (-5.7 to 11.0) | 0.347 |
| Myalgia | 25 (4.4%) | 3 (4.4%) | 22 (4.4%) | 0.1 (-7.7 to 3.7) | 0.971 |
| Sputum production | 51 (9.0%) | 6 (8.7%) | 45 (9.1%) | 0.4 (-8.9 to 5.9) | 0.915 |
| Anorexia | 26 (4.6%) | 6 (8.7%) | 20 (4.0%) | -4.7 (-13.8 to 0.5) | 0.084 |
| Diarrhea | 18 (3.2%) | 4 (5.8%) | 14 (2.8%) | -3.0 (-11.2 to 1.0) | 0.189 |
| Shortness of breath | 26 (4.6%) | 7 (10.1%) | 19 (3.8%) | -6.3 (-15.8 to -0.8) | **0.019** |
| Hemoglobin, g/L | 131 (121 to 143) | 131 (119 to 142) | 132 (121 to 144) | 3 (-2 to 7) | 0.258 |
| Platelet count, $\times 10^9/L$ | 182 (143 to 236) | 163 (138 to 234) | 183 (144 to 237) | 8 (-10 to 25) | 0.390 |
| Blood leukocyte count, $\times 10^9/L$ | 4.8 (3.7 to 6.1) | 5.6 (3.9 to 8.3) | 4.7 (3.6 to 5.9) | -0.9 (-1.6 to -0.3) | **0.004** |
| Neutrophil count, $\times 10^9/L$ | 3.0 (2.2 to 4.2) | 4.4 (2.7 to 6.8) | 2.9 (2.2 to 3.9) | -1.1 (-1.7 to -0.6) | **< 0.001** |
| Lymphocyte count, $\times 10^9/L$ | 1.1 (0.8 to 1.5) | 0.7 (0.5 to 1.1) | 1.2 (0.9 to 1.6) | 0.4 (0.3 to 0.5) | **< 0.001** |
| Alanine aminotransferase, U/L | 20.3 (15.0 to 30.4) | 24.7 (16.8 to 41.0) | 20.0 (15.0 to 29.5) | -3.9 (-7.2 to -0.9) | **0.011** |
| Aspartate aminotransferase, U/L | 24.3 (19.5 to 31.5) | 35.0 (25.6 to 47.0) | 23.8 (19.0 to 29.0) | -10.8 (-14.2 to -7.6) | **< 0.001** |
| Total bilirubin, $\mu\text{mol/L}$ | 11.9 (8.7 to 17.6) | 11.8 (9.0 to 17.4) | 11.9 (8.7 to 17.6) | 0.1 (-1.4 to 1.7) | 0.887 |
| Albumin, g/L | 39.0 (35.7 to 42.4) | 35.4 (31.7 to 38.3) | 39.4 (36.4 to 42.7) | 4.4 (3.2 to 5.7) | **< 0.001** |
| Blood urea nitrogen, mmol/L | 4.0 (3.2 to 5.0) | 4.9 (3.7 to 6.2) | 3.9 (3.1 to 4.8) | -1.0 (-1.4 to -0.6) | **< 0.001** |
| Creatinine, $\mu\text{mol/L}$ | 59.0 (46.8 to 74.0) | 65.1 (53.0 to 84.0) | 58.2 (46.2 to 72.9) | -8.9 (-14.3 to -3.3) | **0.002** |
| Sodium, mmol/L | 137.6 (135.4 to 140.1) | 135.4 (133.3 to 137.2) | 138.0 (135.9 to 140.3) | 2.6 (1.7 to 3.5) | **< 0.001** |
| Potassium, mmol/L | 4.0 (3.6 to 4.3) | 3.9 (3.5 to 4.3) | 4.0 (3.6 to 4.3) | 0.1 (-0.1 to 0.2) | 0.493 |
| Creatine kinase, U/L | 70.8 (48.0 to 112.8) | 81.7 (48.0 to 181.2) | 70.0 (48.0 to 109.3) | -11.4 (-27.0 to 1.7) | 0.092 |
| Lactose dehydrogenase, U/L | 189.0 (152.0 to 244.0) | 304.4 (236.8 to 378.5) | 183.6 (148.3 to 230.3) | -112.2 (-134.0 to -89.9) | **< 0.001** |
| D-dimer $\geq 0.05$ mg/L | 269 (47.7%) | 44 (63.8%) | 225 (45.5%) | -18.3 (-29.5 to -5.7) | **0.004** |
| Procalcitonin $\geq 0.05$ ng/L | 207 (36.7%) | 38 (55.1%) | 169 (34.1%) | -20.9 (-32.8 to -8.5) | **0.001** |
| C-reactive protein level $\geq 10$ mg/L | 286 (50.7%) | 61 (88.4%) | 225 (45.5%) | -43.0 (-50.0 to -32.3) | **< 0.001** |
| Abnormal CT findings | 516 (91.5%) | 68 (98.6%) | 448 (90.5%) | -8.1 (-11.2 to -1.3) | **0.025** |
| Lung CT score | 6 (4 to 11) | 15 (10 to 21) | 6 (3 to 9) | -8 (-10 to -7) | **< 0.001** |

Abbreviations: CI, confidence interval; COPD, chronic obstructive pulmonary disease; COVID-19; 2019 coronavirus disease; CT computed tomography.

**Figure 1.**
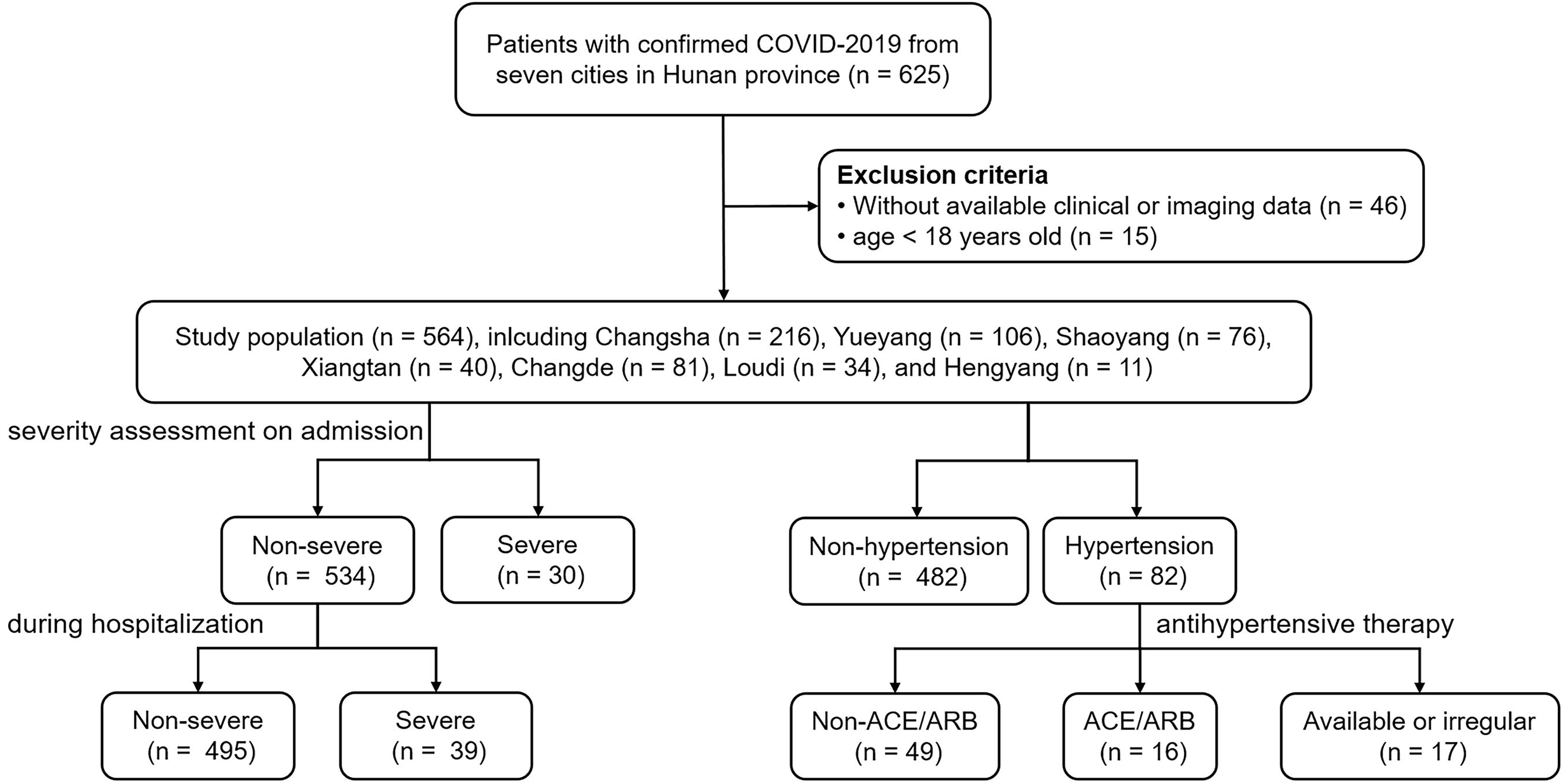
The study design flow chart of the included patients.

### Differences between severe and non-severe patients

In total, 69 of 564 (12.2%) patients were identified to have severe pneumonia. As summarized in Table 1, these patients were older and more likely to have pre-existing comorbidities, including hypertension, diabetes, cardiovascular disease, COPD, and chronic renal disease compared with the non-severe patients. The clinical symptoms at illness onset also differed, with severe patients more likely to present with fever and shortness of breath. Furthermore, patients with severe pneumonia had elevated liver enzyme (aspartate aminotransferase [difference, −10.8; 95% CI, −14.2 to −7.6; *P* < 0.001]), renal dysfunction (blood urea nitrogen [difference, −1.0; 95% CI, −1.4 to −0.6; *P* < 0.001]), abnormal coagulation function (D-dimer ≥ 0.05 mg/L [difference, −18.3%; 95% CI, −29.5% to −5.7%; *P* = 0.004]), raised inflammation-related parameters (lactose dehydrogenase [difference, −112.2; 95% CI, −134.0 to −89.9; *P* < 0.001]), and worse lung CT score (difference, −8; 95% CI, −10 to −7; *P* < 0.001). However, lymphocyte count (difference, 0.4; 95% CI, 0.3 to 0.5; *P* < 0.001) was significantly lower in patients with severe pneumonia.

### Antiviral therapy did not prevent severe pneumonia

Thirty patients were classified with severe pneumonia on admission and were excluded, to leave 534 patients with non-severe COVID-19 on admission for further analysis. Amongst this group, 39 (7.3%) patients developed severe pneumonia during hospitalization. Most (n = 503 [94.2%]) patients received single or combined antiviral therapy during their admission. After adjusting for age, gender, smoking history, hypertension, diabetes, cardiovascular disease, COPD, and lung CT score, none of these investigational antiviral therapies (all *P* > 0.05) were associated with reduced risk of progression to severe disease, as shown in Table 2.

**Table 2.** The impact of different treatment protocols on progression to severe pneumonia after admission (n = 534)

| Protocols | Rude OR (95% CI) | *P* value | Adjusted OR (95% CI) | *P* value |
| --- | --- | --- | --- | --- |
| No antiviral therapy (n = 31) | Reference | 1 | Reference | 1 |
| Arbidol (n = 43) | 1.18 (0.18 to 7.51) | 0.864 | 0.79 (0.11 to 6.01) | 0.823 |
| Lopinavir/ritonavir (n = 71) | 0.91 (0.16 to 5.23) | 0.912 | 0.66 (0.10 to 4.31) | 0.661 |
| Interferon $\alpha$ (n = 18) | 1.81 (0.23 to 14.12) | 0.570 | 1.33 (0.15 to 12.26) | 0.800 |
| Arbidol + Lopinavir/ritonavir (n = 34) | 1.55 (0.24 to 10.01) | 0.643 | 0.95 (0.12 to 7.38) | 0.958 |
| Arbidol + Interferon $\alpha$ (n = 16) | 0.97 (0.08 to 11.54) | 0.979 | 0.23 (0.01 to 4.43) | 0.329 |
| Lopinavir/ritonavir + Interferon $\alpha$ (n = 184) | 1.04 (0.22 to 4.90) | 0.959 | 0.62 (0.12 to 3.35) | 0.579 |
| Arbidol + Lopinavir/ritonavir + Interferon $\alpha$ (n = 137) | 1.45 (0.31 to 6.84) | 0.639 | 0.85 (0.16 to 4.60) | 0.847 |

Abbreviations: CI, confidence interval; OR, odds ratio.
Adjusted OR was calculated by adjusting age, gender, smoking history, hypertension, diabetes, cardiovascular disease, chronic obstructive pulmonary disease, and lung CT score.

### ACEI/ARB therapy in patients with hypertension

Eighty-two individuals were recorded to have hypertension and only 16 patients were on ACEI/ARB therapy (including 6 cases receiving ARB monotherapy, 9 cases receiving combined ARB and calcium-antagonist, and 1 case receiving AECI monotherapy). There were 49 patients on non-ACEI/ARB antihypertensive therapy (including 39 cases receiving calcium-antagonist monotherapy, 3 cases receiving β-blocker monotherapy, 2 cases receiving diuretic monotherapy, and 5 cases receiving combined calcium-antagonist and β-blocker) and all were maintained on their usual antihypertensive regimen during hospitalization. The remaining 17 cases were not on antihypertensive medications or limited by unavailable data and received amlodipine or nitrendipine to control blood pressure as necessary. As summarized in Table 3, hypertensive patients on ACEI/ARB therapy had similar baseline clinical characteristics and laboratory findings compared with those on non-ACEI/ARB antihypertensive therapy. However, patients on ACEI/ARB therapy were less likely to develop severe pneumonia, in contrast with those on non-ACEI/ARB therapy (difference, −26.4%; 95% CI, −41.3%∼-1.5%; *P* = 0.037). Laboratory and imaging data from 65 patients with hypertension during hospitalization were analyzed (Supplementary Figure 2). Lactose dehydrogenase and CT lung scores were lower in patients on ACEI/ARB therapy than those on non-ACEI/ARB therapy at about 3 days after admission.

**Table 3.** Comparison between hypertensive patients on ACEI/ARB therapy and on Non-ACEI/ARB therapy (n = 65)

| Characteristics | ACEI/ARB (n = 16) | Non-ACEI/ARB (n = 49) | Difference (95% CI) | P |
| --- | --- | --- | --- | --- |
| Age (years) | 57 (51 to 65) | 63 (53 to 69) | 5 (-1 to 12) | 0.106 |
| Male gender | 10 (62.5%) | 23 (46.9%) | 15.6 (-11.9 to 38.7) | 0.280 |
| Smoking history | 2 (12.5%) | 5 (10.2%) | 2.3 (-12.4 to 26.5) | 0.797 |
| Comorbidity |  |  |  |  |
| Diabetes | 2 (12.5%) | 18 (36.7%) | 24.2 (-2.2 to 40.9) | 0.068 |
| Cardiovascular disease | 0 (0) | 8 (16.3%) | 16.3 (-4.6 to 29.0) | 0.084 |
| COPD | 0 (0) | 1 (2.0%) | 2.0 (-17.4 to 10.7) | 0.565 |
| Cerebrovascular disease | 0 (0) | 3 (6.1%) | 6.1 (-13.7 to 16.5) | 0.311 |
| Chronic renal disease | 1 (6.3%) | 1 (2.0%) | -4.2 (-26.4 to 5.9) | 0.435 |
| Hepatitis B infection | 0 (0) | 2 (4.1%) | 4.1 (-15.5 to 13.7) | 0.412 |
| Medication history |  |  |  |  |
| Aspirin | 0 (0) | 3 (6.1%) | 6.1 (-13.7 to 16.5) | 0.311 |
| Statins | 1 (6.3%) | 2 (4.1%) | -2.2 (-24.4 to 8.8) | 0.720 |
| Metformin | 1 (6.3%) | 8 (16.3%) | 10.1 (-13.3 to 23.8) | 0.311 |
| Other oral hypoglycemic agents | 1 (6.3%) | 10 (20.4%) | 14.2 (-9.7 to 28.4) | 0.190 |
| Insulin | 0 (0) | 2 (4.1%) | 4.1 (-15.5 to 13.7) | 0.412 |
| Systolic pressure, mmHg | 132 (120 to 137) | 136 (120 to 146) | 7 (-2 to 15) | 0.107 |
| Diastolic pressure, mmHg | 80 (71 to 89) | 80 (78 to 87) | 2 (-3 to 8) | 0.547 |
| Hemoglobin, g/L | 133 (125 to 150) | 133 (118 to 138) | -7 (-17 to 1) | 0.060 |
| Platelet count, $\times 10^9/L$ | 181 (135 to 241) | 182 (147 to 239) | 6 (-38 to 52) | 0.837 |
| Blood leukocyte count, $\times 10^9/L$ | 5.5 (4.6 to 6.5) | 5.2 (3.9 to 7.0) | -0.2 (-1.4 to 0.9) | 0.698 |
| Neutrophil count, $\times 10^9/L$ | 3.7 (3.1 to 4.4) | 3.7 (2.4 to 4.8) | -0.1 (-1.0 to 0.8) | 0.733 |
| Lymphocyte count, $\times 10^9/L$ | 1.1 (0.9 to 1.4) | 1.0 (0.7 to 1.5) | -0.1 (-0.4 to 0.2) | 0.322 |
| Alanine aminotransferase, U/L | 28.9 (16.2 to 40.7) | 23.0 (17.2 to 31.5) | -2.3 (-12.6 to 5.3) | 0.590 |
| Aspartate aminotransferase, U/L | 25.0 (20.0 to 30.9) | 26.3 (20.0 to 36.3) | 0.8 (-4.6 to 7.2) | 0.800 |
| Total bilirubin, $\mu\text{mol/L}$ | 12.6 (10.3 to 19.9) | 12.5 (9.5 to 19.0) | -0.7 (-4.0 to 3.0) | 0.634 |
| Albumin, g/L | 38.4 (34.6 to 44.5) | 37.0 (32.2 to 40.4) | -2.5 (-6.2 to 1.4) | 0.185 |
| Blood urea nitrogen, mmol/L | 4.8 (4.1 to 5.8) | 4.5 (3.7 to 6.5) | -0.2 (-1.2 to 0.9) | 0.611 |
| Creatinine, $\mu\text{mol/L}$ | 64.7 (44.9 to 82.1) | 63.3 (51.2 to 78.7) | 1.3 (-14.5 to 14.7) | 0.877 |
| Sodium, mmol/L | 136.3 (134.7 to 137.2) | 137.5 (134.6 to 140.6) | 1.3 (-0.8 to 3.4) | 0.252 |
| Potassium, mmol/L | 4.0 (3.6 to 4.4) | 3.9 (3.5 to 4.2) | -0.1 (-0.4 to 0.2) | 0.438 |
| Creatine kinase, U/L | 118.5 (64.6 to 187.3) | 64.5 (46.5 to 147.2) | -28.4 (-70.0 to 5.1) | 0.083 |
| Lactose dehydrogenase, U/L | 184.0 (157.1 to 281.7) | 238.0 (164.1 to 315.2) | 33.4 (-19.7 to 89.2) | 0.301 |
| D-dimer $\geq 0.05$ mg/L | 10 (62.5%) | 27 (55.1%) | -7.4 (-30.9 to 19.8) | 0.604 |
| Procalcitonin $\geq 0.05$ ng/L | 6 (37.5%) | 27 (55.1%) | 17.6 (-10.0 to 40.7) | 0.221 |
| C-reactive protein level $\geq 10$ mg/L | 11 (68.8%) | 41 (83.7%) | 14.9 (-6.4 to 40.5) | 0.195 |
| Abnormal CT findings | 15 (93.8%) | 46 (93.9%) | 0.1 (-11.5 to 22.6) | 0.985 |
| CT severity score | 7 (3 to 14) | 10 (5 to 16) | 2 (-3 to 8) | 0.258 |
| Combined treatments after admission |  |  |  |  |
| Nonspecific antiviral therapy | 15 (93.8%) | 45 (91.8%) | -1.9 (-14.1 to 20.7) | 0.803 |
| Interferon $\alpha$ | 8 (50.0%) | 35 (71.4%) | 21.4 (-4.6 to 45.9) | 0.116 |
| Intravenous antibiotics | 12 (75.0%) | 34 (69.4%) | -5.6 (-25.9 to 21.3) | 0.668 |
| Severe pneumonia | 1 (6.3%) | 16 (32.7%) | 26.4 (1.5 to 41.3) | **0.037** |

Abbreviations: ACEI, angiotensin-converting enzyme inhibitors; ARB, angiotensin-receptor blockers; CI, confidence interval; COPD, chronic obstructive pulmonary disease; CT computed tomography.

To further confirm the clinical risk factors for severe pneumonia in COVID-19 patients, univariate and multivariate logistic regression analyses were performed (Table 4). Univariate analyses identified age, hypertension, diabetes, and cardiovascular disease to be associated with the development of severe pneumonia (all *P* < 0.05), but only older age (OR and 95% CI, 1.05 [1.03 to 1.07]; *P* < 0.001) and hypertension without ACEI/ARB therapy (OR and 95% CI, 2.07 [1.07 to 4.00]; *P* = 0.030) were independent risk factors in multivariate analysis. Compared with patients without hypertension, hypertension on ACEI/ARB therapy was not associated with developing severe pneumonia (OR and 95% CI, 0.41 [0.05 to 3.19]; *P* = 0.392).

**Table 4.** Clinical risk factors for severe pneumonia in COVID-19 patients

| Variables | Univariate OR (95% CI) | *P* value | Multivariate OR (95% CI) | *P* value |
| --- | --- | --- | --- | --- |
| Age (per year increase) | 1.06 (1.04 to 1.08) | < 0.001 | 1.05 (1.03 to 1.07) | < 0.001 |
| Male gender | 1.33 (0.80 to 2.20) | 0.275 |  |  |
| Hypertension |  | < 0.001 |  | 0.055 |
| Non-hypertensive | Reference | 1 | Reference | 1 |
| Hypertensive on ACEI/ARB therapy | 0.60 (0.08 to 4.66) | 0.628 | 0.41 (0.05 to 3.19) | 0.392 |
| Hypertensive on other therapy | 3.93 (2.15 to 7.19) | < 0.001 | 2.07 (1.07 to 4.00) | 0.030 |
| Diabetes | 2.95 (1.44 to 6.03) | 0.003 |  | 0.293 |
| Cardiovascular disease | 2.85 (1.08 to 7.56) | 0.035 |  | 0.916 |

Abbreviations: ACEI, angiotensin-converting enzyme inhibitors; ARB, angiotensin-receptor blockers; CI, confidence interval; COVID-19; 2019 coronavirus disease; OR, odds ratio.

### Chloroquine therapy in non-severe patients

Among 534 patients with non-severe COVID-19 on admission, only patients in Changsha received chloroquine therapy (500mg, bid). Thus, 192 non-severe COVID-19 patients without receiving ACEI/ARB therapy in Changsha (25 patients with chloroquine therapy and 167 patients without chloroquine therapy) were included and 25 well-matched pairs for comparison were identified for comparison using PSM, with matched covariates of age, gender, hypertension, and lung CT score (Supplementary Table 1). None of patients treated with chloroquine developed severe pneumonia, though without significance (difference, 12.0%; 95% CI, −3.5% to 30.0%; *P* = 0.074). In addition, there was no significant difference in reported adverse events and ≥ Grade 3 adverse events between the two groups (*P* = 0.777 and 0.440, respectively).

## DISCUSSION

In this study, we described the use of several adjuvant treatments in preventing severe pneumonia development in COVID-19 patients. Hypertensive patients who received ACEI/ARB therapy had a lower risk of developing severe pneumonia compared with those on non-ACEI/ARB antihypertensive therapy, which suggested that ACEI/ARB therapy may be protective from severe pulmonary injury for hypertensive patients. However, these results should be interpreted with caution owing to potential bias and residual confounders in this observational study.

Consistent with earlier reports and clinical experience around the world, our cohort confirmed that that older age and hypertension without ACEI/ARB therapy were independently associated with developing severe pneumonia in COVID-19.^2,3,22^ The association between age and severe pneumonia may be attributed to the accumulation of medical comorbidities with age or decline in immune function over time. Given the close collinearity between age and hypertension, our study adjusted for age by multivariate regression analysis and revealed that hypertension remained an independent risk factor for disease progression irrespective of age for patients without receiving ACEI/ARB therapy.

Our study examined the controversial topic of COVID-19 and ACEI/ARB therapy in hypertensive patients. We found that hypertensive patients on ACEI/ARB therapy were less likely to develop severe pneumonia, although we acknowledged that the analysis was conducted in limited sample size - with only 16 of 82 patients on ACEI/ ARB therapy. The dynamic profile of laboratory and radiological findings further showed that in patient on ACEI/ARB therapy, lactose dehydrogenase and CT lung score were lower in than those on non-ACEI/ARB therapy at about 3 days after admission. Together, these findings were suggestive that ACEI/ ARB therapy may be protective in hypertensive patients with COVID-19.

COVID-19 patients with hypertension may endure worse outcomes due to decreased ACE2 levels. ACE2, an important component of the renin-angiotensin-aldosterone system (RAAS), is highly expressed in both the lung and myocardium and converts angiotensin II (AII) into angiotensin 1-7, which can downregulate ACE and directly cause vasodilatation.^23-25^ Downregulation of ACE2 level has also been shown in hypertensive animal models, although there has been limited success in translational confirmation in human studies.^26,27^ None the less, this may be exacerbated by the further reduction in available ACE2 following coronavirus infection. Both SARS-CoV and SARS-CoV-2 can gain entry into the host cell through binding of the S-protein to the membrane-bound ACE2 aminopeptidase, which then is cleaved by serine proteases to allow for membrane fusion and invasion.^14,28^ Following infection, there is a reduction in pulmonary ACE2, either through internalization with viral entry and/or downregulation of ACE2 enzyme during this process. Hence this may exacerbate the lower baseline ACE2 function in hypertensive patients to lead to the accumulation of AII and reduced angiotensin 1-7 and subsequently worse immune-related cytolysis and lung injury, which was validated by the linear association between elevated plasma AII level and viral load or lung injury in COVID-19 patients.^29^

The use of ACEI or ARB likely promotes feedback upregulation of ACE2 expression in hypertension, although there are animal and human studies which have also found either no effect or downregulation of ACE2 level in the setting of diabetes, hypertension or cardiovascular disease, with or without ACE/ARB therapy.^25,30-33^ Nonetheless, this may explain our finding that ACEI/ARB therapy can be protective in SARS-CoV-2 infection. Further evidence that would support this come from earlier studies in which ACEI/statins were protective in severe pneumonia, recombinant ACE2 protected mice from lung injury following SARS-CoV infection, and RAAS blockade reduced acute lung injury in rats injected with SARS-CoV S-protein. ^34,35^

Currently, there is still no specific treatment for COVID-19 except for supportive care. In our cohort, 94.2% of non-severe patients received nonspecial antiviral therapy from just after admission. The combination protocol and medication dose varied in different hospitals. However, our results did not support any single or combined antiviral treatments to limit the progression of COVID-19. In addition, none of 25 patients treated with chloroquine developed severe pneumonia, though without statistical significance. This is unfortunately limited by the degree of heterogeneity in our cohort and relatively small size. Chloroquine is known to block virus infection by increasing endosomal pH required for virus/cell fusion, as well as interfering with the glycosylation of cellular receptors of coronavirus, which has been confirmed to inhibit SARS-CoV-2 in vitro.^36-38^ A recent open□label non□randomized clinical trial reported that hydroxychloroquine combined with azithromycin is significantly associated with viral load reduction/disappearance in COVID-19 patients. ^39^ Even so, more RCTs to assess the therapeutic effects of chloroquine on COVID-19 need to be conducted.

## LIMITATIONS

There were several limitations in this study. First, our study was conducted by retrospective data analysis. Second, some cases had incomplete laboratory test results, given the variation in the clinical management among different hospitals. All data generation was clinically driven and not systematic. Third, because most patients had recovered and discharged at the time of data cutoff and only 2 patients died during admission, the impact of adjuvant treatment on the clinical outcomes cannot be analyzed. Forth, there were limited hypertensive patients who received ACEI or ARB therapy, so we cannot analyze the effect of AECI and ARB respectively. A larger cohort study of COVID-19 patients with hypertension from other cities in China and other countries would help to further validate the potential protective effect of AECI/ARB therapy.

## CONCLUSIONS

COVID-19 is a fast-evolving, global pandemic with high mortality from severe pneumonia, ARDS, and multiorgan failure, with the highest risk in the elderly and patients with underlying hypertension. The current adjuvant antiviral and immunoregulation therapies have not yet proven benefits in clinical settings. But importantly, our results show the potential protective effects of ACEI or ARB therapy to limiting progression to severe lung disease in hypertensive patients.

## Data Availability

All data referred to in the manuscript need to be requested to the corresponding authors.

## ACKNOWLEDGEMENT

We thank Hongzhuan Tan, PhD, from the Department of Epidemiology, Xiangya School of Public Health, Central South University for his statistical analysis discussion. We also thank Wei Nie, MD, Qin Liu, MD, Jing Zhao, MD, Junhong Duan, MD, Zhimin Yan, MD, Min Yang, MD from Department of Radiology, Ying Li, MD from Health Management Center, Sujie Jia, MD from Department of Pharmacy, Third Xiangya Hospital, Central South University, for their data collection. They were not compensated for their contributions.

## Author Contributions

Drs W. Wang and P. Rong had full access to all of the data in the study and take responsibility for the integrity of the data and the accuracy of the data analysis. Drs P. Rong and W. Wang contributed equally to this article.

Concept and design: Z. Feng, J. Li, S. Yao, X. Ma, P. Rong, W. Wang; Acquisition, analysis, or interpretation of data: Z. Feng, Q. Yu, W. Zhou, X. Mao, H. Li, W. Kang, X. Ouyang, J. Mei, Q. Zeng, J. Liu; Drafting of the manuscript: Z. Feng, J. Li, S. Yao; Critical revision of the manuscript for important intellectual content: J. Li, X. Ma, P. Rong, W. Wang; Statistical analysis: Z. Feng, S. Yao, W. Kang; Obtained funding: P. Rong; Administrative, technical, or material support: S. Yao, Q. Yu, W. Zhou, X. Mao, X. Ma, P. Rong, W. Wang.

